# Soluble suppression of tumorigenicity-2 (sST2) predicts mortality and right heart failure in LVAD patients

**DOI:** 10.1101/2023.02.06.23285564

**Authors:** Lieke Numan, Emmeke Aarts, Faiz Ramjankhan, Marish I.F. Oerlemans, Manon G. van der Meer, Nicolaas de Jonge, Anne-Marie Oppelaar, Hans Kemperman, Folkert W. Asselbergs, Linda W. Van Laake

## Abstract

**Background:** Soluble suppression of tumorigenicity-2 (sST2) predicts mortality in heart failure patients. The predictive value of sST2 in left ventricular assist device (LVAD) patients remains unknown. Therefore, we studied the relationship between sST2 and outcome after LVAD implantation.

**Methods:** Patients implanted between January 2015 and December 2022 were included. Survival of patients with normal and elevated pre-operative sST2 levels was compared using Kaplan-Meier analysis. The relationship between post-operative sST2, survival and right heart failure (RHF) was evaluated using a Joint Model (JM). Multivariate JM analysis adjusted for serially measured NT-proBNP was performed.

**Results:** The median follow-up was 25 months, during which 1573 post-operative sST2 levels were measured in 199 patients, with a median of 29 ng/ml. Survival in patients with normal or elevated pre-operative sST2 levels (n=86) did not differ significantly (p=0.22). Time-dependent post-operative sST2 levels were significantly associated with mortality, with a hazard ratio (HR) of 1.20 (95% CI: 1.10-1.30, p<0.01) and a HR of 1.22 (95% CI: 1.07-1.39, p=0.01) for RHF, both per 10 unit sST2 increase. The sST2 instantaneous change was not predictive for survival or RHF (p=0.99, p=0.94 respectively). Multivariate JM analysis showed a significant relationship between sST2 with mortality adjusted for NT-proBNP with HR 1.19 (95% CI: 1.00-1.42, p=0.05), while the HR of RHF was not significant (1.22, 95% CI: 0.94-1.59, p=0.14), both per 10 unit sST2 increase.

**Conclusion:** Time-dependent post-operative sST2 predicts all-cause mortality after LVAD implantation independently of NT-proBNP. Future research is warranted into possible target interventions and the optimal monitoring frequency.

## Introduction

Soluble suppression of tumorigenicity-2 (sST2) is a prognostic indicator for mortality in heart failure (HF) patients and may help in risk stratification in HF patients.(1) Although the mechanism has not been elucidated completely, it is known that sST2 is part of the ST2/interleukin-33 (IL33)-pathway, which is activated by increased cardiac wall stress.(2) Cardiac myocytes and lung epithelial cells are amongst the sources of sST2 production, enhanced by pro-inflammatory cytokines.(3)(4) Normally, IL-33 binds to the membrane bound transmembrane isoform of ST2 (ST2L) and thereby prevents apoptosis and fibrosis. Soluble ST2 acts as a decoy receptor for IL-33, so the cardioprotective effect of IL-33 is lost when IL-33 binds to soluble ST2.(5)

In contrast to the general HF population, only a few studies examined the role of sST2 in patients with end-stage heart failure. Left ventricular assist device (LVAD) implantation results in unloading of the left ventricle and promotes reverse modeling(6). Despite improving survival and complication rates, patients on LVAD support have a relatively high mortality rate and frequently suffer from adverse events such as right heart failure. (7)(8) sST2 could contribute to stratification and early recognition of deterioration in patients on LVAD support. Tseng et al. demonstrated very high sST2 levels in patients with end-stage HF before LVAD implantation, especially in patients in cardiogenic shock at implantation. After LVAD implantation, sST2 levels dropped significantly, with normalized values after three months, suggesting that LVAD implantation leads to a reduction of fibrosis and inflammation(9). Opfermann et al. showed a significant increase in sST2 during the initial post-operative period after LVAD implantation and normalization to pre-operative levels after one week and normal levels after three weeks. (10)

Although repeated sST2 measurements appeared to be a strong predictor of outcome in patients with acute HF(11), the predictive value of serially measured sST2 levels in LVAD patients on long term outcomes has not been elucidated yet. Therefore, we aimed to assess the relationship of serially measured sST2 and long-term outcome in patients on LVAD support. We hypothesized that elevated or increasing sST2 levels predict adverse outcome such as right heart failure or death.

## Methods

### Study design

In this single Center retrospective cohort study patients receiving a HeartWare (HVAD) or HeartMate3 (HM3) between January 2015 until December 2021 in the University Medical Center Utrecht (UMCU) were included. The follow-up was until May 2022.

The study was approved by the local ethics committee of the UMCU (no: 20-195) and was handled in accordance with the Declarations of Helsinki and Good Clinical Practice. The need for informed consent for the use of retrospective data was waived. Data were retrieved from the electronic health records.

### Endpoints

The primary endpoint of the study was death or urgent heart transplantation (HTx) during follow-up. Transplantation was labelled as urgent HTx if a patient had received priority status on the waiting list (National 1A, national 1B, or international HU). Without the urgent heart transplantation, the patient would probably not have survived and we therefore considered this as adverse outcome in addition to death. Patients were censored for ongoing support at the end of follow-up, explantation and non-urgent HTx. The secondary endpoint was right heart failure, which was defined using the definitions of the Interagency Registry for Mechanically Assisted Circulatory Support (INTERMACS) as: requiring right ventricular assist device support or nitric oxide inhalation and/or inotropic therapy for more than 1 week at any time after LVAD implantation. For the current study we included the first right heart failure event > 30 days post-implantation for analysis, since sST2 was mostly measured after the initial post-operative period.

### sST2 measurement

Both pre- and post-operative sST2 levels were analyzed. sST2 was measured during regular outpatient visits (every 3-4 months) and from biobanked samples if available. sST2 was measured in heparin plasma using the ASPECT-PLUS ST2 assay on an ASPECT Reader (Critical Diagnostics, San Diego, CA, USA). Biobanked samples were centrifuged within 6 hours after withdrawal, and plasma was stored at −80°C. Levels of sST2 using biobanked samples after a maximum of one freeze/thaws cycle were analyzed by using the Presage ST2 ELISA assay according to manufacturer’s instructions (Critical diagnostics, San Diego, CA, USA).(9) The ASPECT Reader and Presage ST2 ELISA techniques are comparable (R^2^=0.92). (12)

### Statistics

#### Pre-operative sST2

We assessed the primary endpoint in patients within normal pre-operative sST2 limits (≤35 ng/ml) compared to patients with elevated pre-operative sST2 (>35 ng/ml, which is the current cut-off value used for heart failure patients(13)) using Kaplan-Meier analysis. The last sST2 measurement within 30 days before the primary implantation was included.

#### Post-operative sST2

The intracluster correlation coefficient was calculated to express the variation of post-operative sST2 within patients and between patients. To visualize the trajectory of post-operative sST2, a heatmap was created, which is a visualization technique (not for quantification) that shows the height of the average sST2 levels for all patients in one figure. Time bins of 90 days were used, and the heatmap was sorted on patients with the highest sST2 measurement in the entire dataset. Primary and secondary endpoints were indicated in the heatmap.

In addition, to study the association between longitudinal sST2 measurements and our primary and secondary endpoints using a joint model (JM) approach(14). A JM approach was chosen, since it deals with irregular measurement intervals and missing data and was used in comparable settings in previous studies. (15)(16) Within this two-step method, the trajectory of sST2 over time is combined with a time-to-event model. The longitudinal pattern of sST2 is estimated using linear mixed effects (LME) modeling). The LME included a random effect for both the intercept and time, with patient ID as the clustering level. A natural spline was used to allow for non-linearity in the association between time and sST2, with a knot at 90 days. Four random cases were displayed, to visualize the estimated trajectory and the sST2 measurements. Time to-event analysis in the JM was done using two cause-specific multivariable Cox regression for the primary and secondary outcome, stratified for device type (HVAD or HM3) adjusting for clinically relevant variables: age at primary implantation, body mass index (BMI), sex, temporary mechanical support at primary implantation, INTERMACS, ischemic etiology and right ventricle (RV) function. RV function was classified as poor, moderate or good by two independent cardiologists, who individually classified the RV based on echocardiography and right heart catheterization using earlier published methods.(17) Within the JM, both the effect of the predicted sST2 and the predicted change in sST2 at the time of the event (the instantaneous slope) on the primary and secondary outcome were assessed. As a sensitivity analysis, a multivariate JM analysis, was performed, adjusted for serially measured NT-proBNP, with a log_10_ transformation of NT-proBNP and a natural spline with a knot at 90 days. In this multivariate JM similar covariates as in the primary analysis were adjusted for.

In literature, patients that receive a heart transplantation, including patients that were listed as high urgent, are usually censored. Hence, we performed a sensitivity analysis similar to the primary JM analysis, now also censoring for urgent HTx.

To assess whether post-operative sST2 trajectories vary in patients on different types of LVAD support (HVAD, HM3), we fitted another separate linear mixed model LME model similarly to the LME model in the JM analysis, and added device type as a fixed variable. Significance of the prediction of the sST2 trajectory by device type was assessed using a Chi^2^-log likelihood test comparing models with and without the predictor device type.

#### Pre- and post-operative sST2

To assess the correlation between pre-operative sST2 and post-operative sST2, pre-operative sST2 levels and predicted sST2 levels at the end of the follow-up were visualized in a scatterplot. Moreover, to quantify this relationship, the Spearman correlation coefficient of the pre-operative sST2 and predicted sST2 at the end of the follow-up or at the time of the event was calculated in all patients that were included in the primary JM analysis and of whom pre-operative sST2 was available.

The flowchart of patients that were included for each of the analysis is displayed in the graphical abstract. Model assumptions of both linear mixed models and Cox regression were checked. P-values < 0.05 were considered significant. Results are presented as median and inter-quartile range (IQR) for continuous variables and as number or percentages for categorical variables. All analyses were performed using R version 3.6.3.

## Results

Between 2015 and January 2022 237 patients received either HVAD or HM3 as a primary LVAD implantation at the University Medical Center Utrecht. Baseline characteristics of all patients with and without post-operative sST2 measurement(s) are presented in Table 1, with complete covariates for all patients. The median age at implantation was 56 years (IQR: 16 years) and 65% of the patients were male. Most patients had non-ischemic cardiomyopathy as underlying disease (70%). The median follow-up time was 25 months (IQR: 33 months), during which 61 (26%) patients died after a median of 10 months (range: 0-69 months). Table 2 shows the causes of death. 8 patients (3%) received urgent HTx after a median of 26 months (range: 1-66 months), 15 patients (6%) received normal HTx after a median of 36 months (range: 11-64) and no patients were weaned from LVAD support. Right heart failure > 30 days after surgery was diagnosed in 26 patients (11%). None of these patients were treated by a right ventricular assist device (RVAD) implant.

**Table 1:**
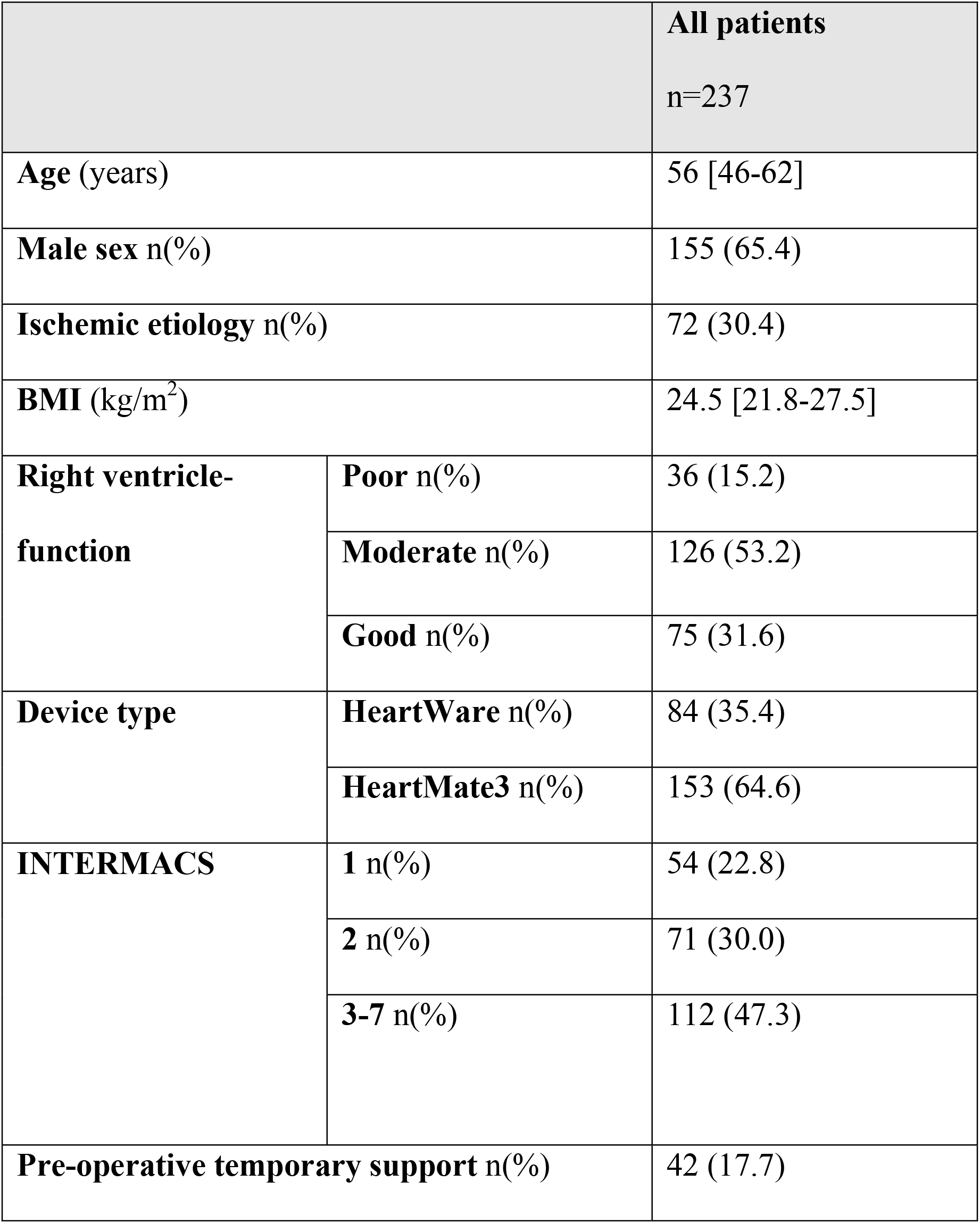
Baseline characteristics of all patients (n=237).

**Table 2:** Causes of death of all patients.

| Type of death | Number of patients n<br>(%), total = 61 |
| --- | --- |
| Device malfunction | 3 (4.9) |
| Infection | 10 (16.4) |
| Multi-organ failure | 12 (19.7) |
| Neurological | 13 (21.3) |
| Right heart failure | 9 (14.8) |
| Other | 14 (23.0) |

### Pre-operative sST2

sST2 was measured pre-operatively (<30 days before surgery) in 86 patients, with a median of 57 ng/ml (IQR: 57 ng/ml). Figure S1 depicts a boxplot of the pre-operative sST2 in patients that reached the primary endpoint and patients that did not. Supplemental table S1 shows the baseline characteristics of patients with and without pre-operative sST2. Patients with pre-operative sST2 level more often had a good RV-function, were more frequently implanted with a HM3, were less often on temporary mechanical support and were more often classified as INTERMACS 3-7.

Figure 1 depicts the Kaplan-Meier survival of patients with high (>35 ng/ml in 64 patients) or normal (≤35 ng/ml in 21 patients) pre-operative sST2 levels. Survival of patients with normal or elevated pre-operative sST2 levels did not differ significantly (p=0.22).

**Figure 1:**
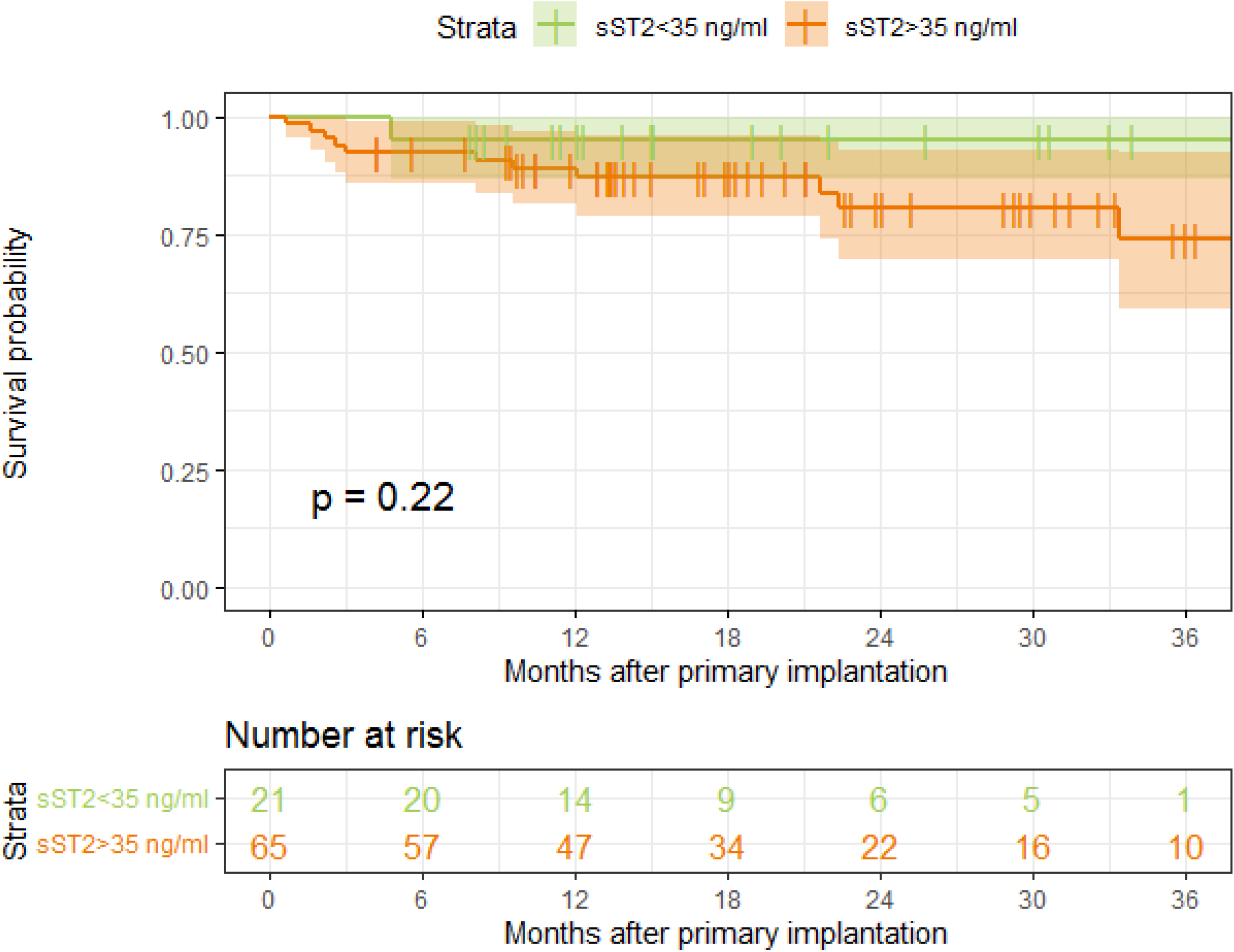
Kaplan–Meier survival of patients with a normal or elevated pre-operative soluble suppression of tumorigenicity-2 (sST2) level.

### Post-operatives sST2

In total, 1573 post-operative sST2 levels were measured in 199 patients, with a median of 29 ng/ml (IQR: 21 ng/ml). 38 patients out of 237 had no post-operative sST2 measurements, of which the majority died early. Figure 2 depicts the post-operative sST2 measurement and its predicted trajectories of four random cases. All predicted sST2 trajectories are displayed in Figure S2, separated for patients with and without event (death, urgent HTx, right heart failure or no event). The intracluster correlation coefficient of post-operative sST2 was 0.73, so 73% of the variation observed in post-operative sST2 is explained by differences between patients, and the remaining 27% is a result of variation within patients over time.

**Figure 2:**
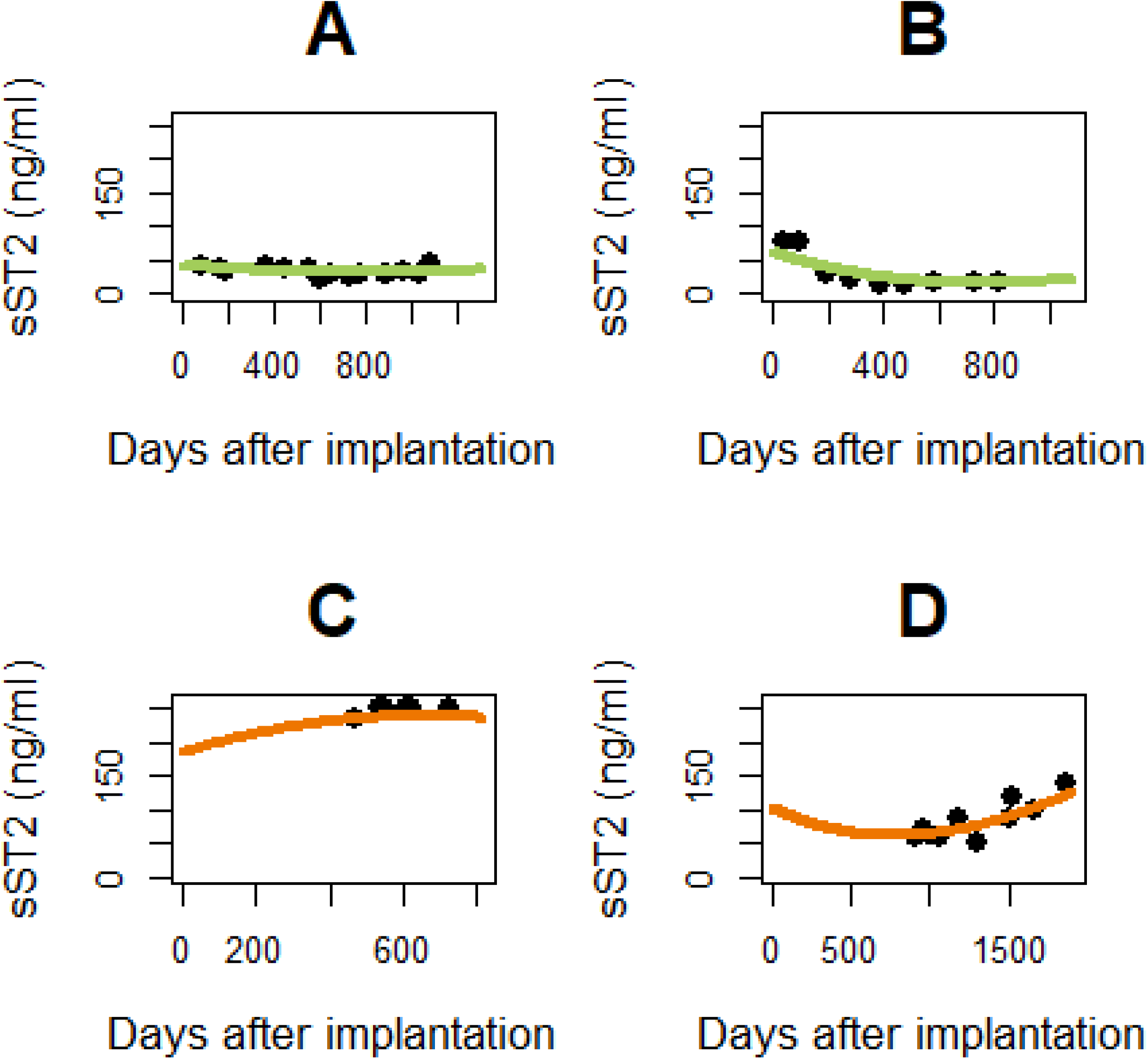
Trajectory of post-operative soluble suppression of tumorigenicity-2 (sST2) levels in four patients. The black dots indicate the sST2 measurement, and the predicted sST2 trajectory by the linear mixed effects model is depicted by the colored line for patients (A and B) without event (A and B) or patients that reached the primary endpoint (C and D).

Figure 3 shows a heatmap sorted by highest sST2 level in the entire post-operative sST2 measurement set, visualizing both primary and secondary endpoints and the height of the sST2 level for all patients. The majority of the patients that reached the primary (survival) and secondary outcome (right heart failure) are concentrated in the lower part of the figure, with higher sST2 levels over time, suggesting that patients that reached the primary outcome have higher sST2 values. To test this finding, Joint Model (JM) analysis was performed, which confirmed the predictive value of time-dependent sST2 for survival with HR = 1.20 (95% CI: 1.10-1.30, p<0.01) for a 10 unit increase in sST2 (Table 3). In addition, time dependent sST2 was a predictor for right heart failure as well, with a HR of 1.22 (95% CI: 1.07-1.39, p=0.01) for a 10 unit increase. A sensitivity analysis where we censored for urgent transplantation, instead of considering it a proxy for mortality, confirmed the significant relation between time-dependent sST2 and survival, with a HR of 1.18 (95% CI: 1.08-1.30 p<0.01) for a 10 unit increase of sST2. Thus, higher expected levels of sST2 at a point in time are predictive for a worse survival. We also investigated whether the instantaneous dynamics sST2 values at the time of the events would be more predictive than the time-dependent predicted values. However, the predicted instantaneous change in sST2 at the time of the event was not an independent predictor of survival (HR: 0.99 (95% CI: <0.01 - >10.0, p=0.99) nor of right heart failure (HR: 1.15 95% CI: <0.01 - >10.0, p=0.94).

**Table 3:** Joint model results for soluble suppression of time-dependent tumorigenicity-2 (sST2) and survival and right heart failure.

|  | Survival |  | Right heart failure |  |
| --- | --- | --- | --- | --- |
|  | Hazard Ratio<br>[95% CI] | p-value | Hazard Ratio<br>[95% CI] | p-value |
| Age at implantation | 0.99 [0.96-1.02] | 0.58 | 1.00 [0.96-1.03] | 0.83 |
| Sex (1=male) | 1.32 [0.59-3.07] | 0.53 | 1.30 [0.43-4.29] | 0.69 |
| Ischemic etiology | 1.13 [0.46-2.89] | 0.83 | 0.52 [0.11-1.90] | 0.37 |
| BMI (kg/m<sup>2</sup>) | 1.06 [0.98-1.13] | 0.11 | 0.97 [0.87-1.08] | 0.57 |
| RV-function | 0.78 [0.46-1.32] | 0.36 | 1.18 [0.53-1.39] | 0.71 |
| INTERMACS | 1.02 [0.67-1.50] | 0.92 | 0.52 [0.26-1.01] | 0.06 |
| Time-dependent sST2 | 1.20 [1.10-1.30] | <0.01 | 1.22 [1.07-1.39] | 0.01 |

CI: confidence interval, BMI: body mass index, RV: right ventricle.

**Figure 3:**
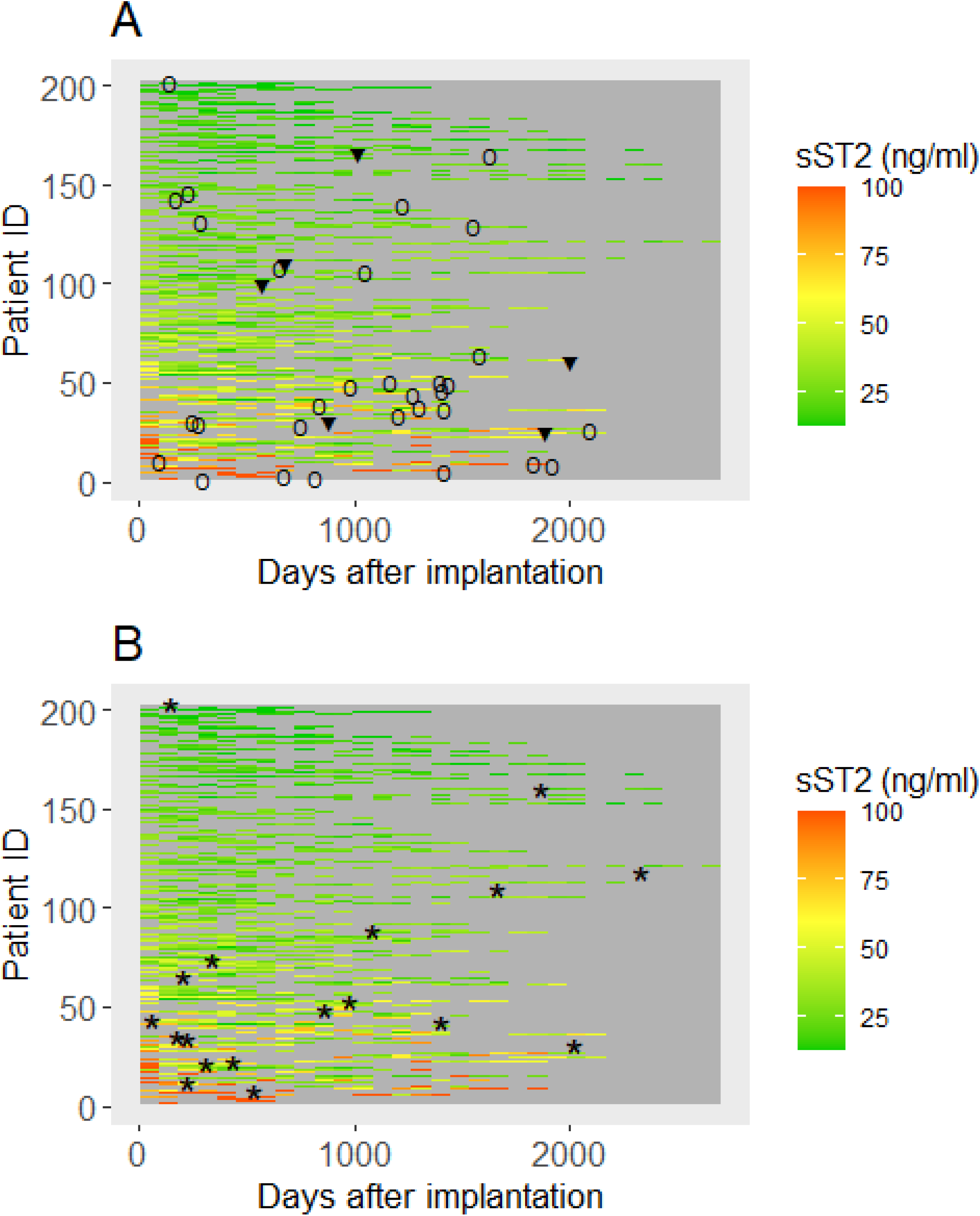
Heatmap for death or urgent heart transplantation (A) and right heart failure (B) of all post-operative soluble suppression of tumorigenicity-2 (sST2) levels, sorted by highest value. The triangle (▼) indicates urgent heart transplantation, the circle (Ο) indicates death and the asterisk (*) indicates right heart failure.

Multivariate JM analysis, adjusted for serially measured NT-proBNP, showed a significant relationship with mortality and urgent HTx with HR 1.19 (95% CI: 1.00-1.42, p=0.05) per 10 unit increase of time-dependent sST2. As an example, this means that the HR is 1.19 for an sST2 of 30 instead of 20 ng/ml sST2. For RHF the relationship was not significant after adjustment for NT-proBNP, with HR 1.22 (95% CI: 0.94-1.59, p=0.14) per 10 unit increase. In the multivariate JM, the HR of time-dependent NT-proBNP for mortality was 3.22 (95% CI: 0.62-16.5, p=0.14) for a NT-proBNP level that is 10 times higher. As an example, this implies a HR of 3.22 for an NTproBNP of 2000 instead of 200 pg/ml. For RHF, the HR of time-dependent NT-proBNP was 2.18 (95% CI: 0.28-14.2, p=0.42) for a NT-pro-BNP level that is 10 times higher in the multivariate JM.

In addition, we explored the potential role of device type as a possible confounder for survival.(17)(18) In a sensitivity analysis using LME with device type as additional fixed effect, device type was not a significant predictor of sST2 at the primary outcome or at the end of the follow-up (p=0.26).

Finally we studied the relation between pre-operative sST2 and predicted post-operative sST2 at the event or end of the follow-up (figure S3). The Spearman correlation coefficient of pre-operative sST2 and predicted post-operative sST2 at the end of the follow-up was 0.05 (p=0.63).

## Discussion

In this study on long term serial sST2 measurements in a large group of patients on LVAD support, we demonstrated that time-dependent sST2 levels after LVAD implantation are predictive for mortality and RHF. The time-dependent sST2 level was predictive for both right heart failure and mortality, whereas the instantaneous increase of sST2 at the event or end of the follow-up was not a significant predictor. In addition, time-dependent sST2 is a significant predictor for mortality independently of time-dependent NT-proBNP (graphical abstract). sST2 trajectories did not differ between patients on different types of LVAD support. We found no significant difference in survival on LVAD in patients with and without elevated pre-operative sST2 levels.

sST2 has been studied extensively in general heart failure patients for more than a decade(5). Vark et al. studied the prognostic value of serial sST2 measurements in patients with acute heart failure and suggested that an increase or stabilization of sST2 levels may be useful in daily practice for stratification or to monitor treatment.(11) Moreover, sST2 is an independent risk factor for hypertensive left ventricular hypertrophy.(19) Repeated sST2 measurements provide independent prognostic information in addition to NT-proBNP in patients with acute heart failure. As NT-proBNP is a biomarker indicating volume overload, while sST2 is a marker of remodeling, inflammation and cardiac fibrosis, the two biomarkers are complementary. (11,20,21) sST2 levels may be increased in other, mostly inflammatory-associated diseases such as COPD, pneumonia or sepsis. (22) However, in contrast to NT-proBNP, sST2 is independent of important prognostic factors such as age, sex, BMI, hypertension, smoking, prior myocardial infarction or renal dysfunction.(23)(24)(25)(26) and only modestly positively correlated with NT-proBNP.(24) In a systematic review, Janssen et al. concluded that natriuretic peptides before LVAD implantation are not predictive of all-cause mortality.(27) On the other hand, Bellavia et al. showed that higher pre-operative levels of NT-proBNP were associated with a higher risk of post-LVAD RHF.

Recently, some short-term studies on sST2 were performed in patients with end-stage heart failure on LVAD support[9], [26]. In a prior pilot study, we reported sST2 analysis in 38 patients with end-stage heart failure both before and up until 6 months after LVAD implantation. sST2 was elevated in patients just before LVAD implantation and decreased to normal values within three to six months after primary LVAD surgery. No difference was observed between male and female patients.(9) In contrast, Denfeld et al. found a different sST2 trajectory in 79 male and 19 female patients within the first 6 months after LVAD implantation, whereas fairly similar trajectories in symptoms were demonstrated from pre- to 6 months post-implantation. Female patients showed an initial increase after implantation, followed by a decrease, whereas male patients showed an overall decrease.(28) In the current study, none of the covariates including sex were independent predictors for adverse outcome in addition to longitudinal sST2 measurements.

Our study revealed no significant relation between the instantaneous slope of sST2 and neither primary or secondary outcome. This is in line with the findings of Vark et al. who found no significant relationship between the instantaneous slope of sST2 and all-cause mortality and HF rehospitalization in HF patients. (11) However, the relationship may be diminished due to the limited monitoring frequency or relatively low number of patients with increasing levels of sST2, which therefore might have led to an underestimation of the effect slope by the linear mixed model.

To the best of our knowledge, we were the first to evaluate the predictive value of serially measured sST2 in patients on long-term LVAD support independently of NT-proBNP. For the current study we used a JM approach, to allow for analysis with irregular and longitudinal sST2 measurements and different types of outcome. JM allows for individual variation, due to the use of a random effect for both the intercept and time in the linear mixed effect models. (14) In addition, it allows for evaluation of the effect of an instantaneous slope in the biomarker of interest in addition to the predicted instantaneous value at the time of the event (or end of the follow-up). To deal with the competing risks, we used two cause-specific Cox models in the JMs. The cause-specific hazard function indicates the hazard rate of an event in subjects who are currently free of events. (29) Within JM we were also able to adjust for important covariates, including serially measured NT-proBNP.

### Limitations

This study has some limitations. sST2 was not measured at fixed intervals in all patients from the start in 2015. Therefore sST2 measurements in some patients were irregular or absent. Pre-operative sST2 levels were present in a relatively small subset of patients (86/237), including patients that were more stable at the time of implant, regarding INTERMACS and RV-function. Therefore, this cannot directly be extrapolated to the whole LVAD population. To deal with missing and irregular data, we used a JM approach that estimates the trajectory of post-operative sST2 levels. Due to the relatively low number of events, we were not able to distinguish predictive values specific for different types of cause of deaths. Possibly, the association between time-dependent sST2 and RHF after adjustment for serially measured NT-proBNP did not reach statistical significance due to a relatively small patient cohort. The majority of the population was Caucasian and therefore the results cannot be extrapolated to other ethnicities Lastly, due the maximum number of covariates in the JM is related to sample size. Increasing patient numbers would allow for additional clinically relevant parameters, such as early post-operative heart rate or the diurnal pattern of sST2. (30)(31)

### Future

Now that the predictive role of sST2 both early and late after LVAD implantation has been established, the question rises how often and at which moments sST2 should be measured. Since high frequency monitoring results in high costs and patient burden, future research is warranted to define the optimal monitoring frequency. Repeated echocardiography and pump parameter evaluation is suggested for patients with persistently high sST2. In addition, it is recommended to investigate whether sST2 responds to interventions such as heart failure medication or pump speed adjustment. Also, future studies with higher frequency sST2 measurements in a larger patient cohort should define a cut-off value for LVAD patients, being a specific subgroup of HF patients. The current study focused on repeatedly measured sST2 and identified it as a crucial biomarker, independently of NT-proBNP, but prediction models can be enhanced in the future by adding other biomarkers or clinical and echocardiographic variables. Altogether, early interventions in LVAD patients with elevated sST2 levels may assist in preventing complications.

In conclusion, sST2 predicts all-cause mortality and right heart failure after LVAD implantation. Moreover, sST2 is a predictor for mortality independently of NT-proBNP. The time-dependent level of post-operative sST2 is predictive, in contrast to the instantaneous change in sST2 at the time of the event or end of the follow-up. sST2 trajectories after LVAD implantation do not cluster per type of LVAD. A closer follow-up can be appropriate in patients with high sST2 levels. Further research is warranted into the optimal monitoring frequency, the mechanisms of elevated sST2 in LVAD patients and possible targeted interventions.

## Data Availability

Data will not be made publicly available.

## Non-standard Abbreviations and Acronyms

HM3: HeartMate 3
HVAD: HeartWare
HTx: heart transplantation
IL-33: interleukin-33
INTERMACS: Interagency Registry for Mechanically Assisted Circulatory Support
JM: joint model
LME: linear mixed effects
LVAD: left ventricular assist device
sST2: Soluble suppression of tumorigenicity-2
UMCU: University Medical Hospital Utrecht

## Figure legends

**Central illustration:** Flow chart showing the patient flow into each analysis: pre-operative and post-operative soluble suppression of tumorigenicity-2 (sST2) measurement(s).

**Supplemental Figure 1:** Pre-operative soluble suppression of tumorigenicity-2 (sST2) of patients that died or underwent urgent heart transplantation and patients that were censored.

**Supplemental Figure 2:** Predicted post-operative individual soluble suppression of tumorigenicity-2 (sST2) trajectory of patients that died or received urgent heart transplantation (HTx) in (A), patients suffering from right heart failure in (B) and patients without primary or secondary outcome in (C). The “normal” cut-off value (35 ng/ml) is indicated by the dashed black line, and the smoothened sST2 trajectory is depicted in grey.

**Supplemental Figure 3:** Relation between pre-operative soluble suppression of tumorigenicity-2 (sST2) and predicted post-operative sST2 at the event or end of follow-up categorized for device type.

